# Do predictors of adherence to pandemic guidelines change over time? A panel study of 21,000 UK adults during the COVID-19 pandemic

**DOI:** 10.1101/2020.11.10.20228403

**Authors:** Liam Wright, Daisy Fancourt

## Abstract

In the absence of a vaccine, governments have focused on behaviour change (e.g. social distancing and enhanced hygiene procedures) to tackle the COVID-19 pandemic. Existing research on the predictors of compliance with pandemic measures has often produced discrepant results. One explanation for this may be that the determinants of compliance are context specific. Understanding whether this is the case is important for designing public health messaging and for evaluating the generalisability of existing research. We used data from the UCL COVID-19 Social Study; a large weekly panel of UK adults from first five months of lockdown in the UK (n = 21,000). We tested whether the extent to which demographic, socio-economic position, personality traits, pro-social motivations, and the living environment predict compliance changed across the pandemic. Low compliance was strongly related to younger age and also to risk attitudes, empathic concern, and high income, among other factors. But the size of some of these associations was larger in later months when less stringent lockdown and household mixing measures were in place, suggesting context-specific effects. The results also showed that compliance fell faster across some groups, suggesting the importance that public health communications adopt a plurality of messages to maximize broad adherence.

## Introduction

Governments have implemented a range of measures to tackle the COVID-19 pandemic. In the absence of a vaccine, measures have focused on reducing transmission of the virus through isolating those with diagnosed or suspected COVID-19, increasing ‘social distancing’ (e.g. ‘shelter-at-home’ orders, restricting non-essential travel and limiting groups gathering in public venues), and enhancing hygiene procedures (such as the wearing of face masks). These measures has been shown to reduce infection and, consequently, lower overall mortality rates (Chu et al., 2020; Mongey et al., 2020). However, each of these measures requires citizens to make voluntary changes to their usual behaviour, sometimes at considerable personal cost (Adams-Prassl et al., 2020). Ensuring high levels of compliance has been a considerable challenge (Fancourt et al., 2020; Park et al., 2020; Wang et al., 2020). Therefore, understanding the factors that determine compliance is vital for managing the pandemic.

There is a sizeable literature on the determinants of compliance with social distancing, hygiene, and quarantine rules, both from the COVID-19 pandemic and from previous epidemics (for reviews, see Bish & Michie, 2010; Webster et al., 2020). The literature highlights the importance of socio-economic and demographic characteristics (e.g. age, gender, education and working status), personality traits (e.g. Big-5 traits and self-efficacy), pro-social motivations (e.g. social capital and empathic concern), and the lived environment (such as household overcrowding and availability of green space), in predicting compliance levels. The results in this literature are not always consistent, however. For instance, younger people, males, and more extroverted individuals are typically found to have lower compliance than other individuals, but some studies show no effects (Bish & Michie, 2010; Clark et al., 2020; Zajenkowski et al., 2020).

One explanation for these discrepancies may be that the determinants of compliance are context-specific. For instance, self-efficacy beliefs, old age, and risk-preferences may change in importance as cases rise or as external restrictions on behavior change. This is consistent with models of human behavior such as the “COM-B” framework, which posits that capabilities, opportunities and motivations combine to determine behavior (Michie et al., 2011). For example, low self-efficacy may undermine psychological *capability* and risk-preferences and age may factor into non-compliance *motivations*, but strict lockdown may limit *opportunities* for non-compliance. Strict lockdown may thus represent a “strong situation” where opportunities for non-compliance are restricted and the information regarding desired behavior is clear, making personality and other characteristics less important for compliance behaviour (Götz et al., 2020; Zajenkowski et al., 2020).

This argument is supported by findings from the H1N1 pandemic that the predictors of compliance differed through time (van der Weerd et al., 2011). It is also supported by findings from the COVID-19 pandemic that the Big-5 personality traits neuroticism and openness were more highly related to compliance with “shelter-at-home” guidelines in areas where less stringent measures were in place (Götz et al., 2020). Overall compliance levels do not remain stable across pandemics (Cowling et al., 2010; YouGov, 2020), which also raises the possibility that the composition of non-complying groups varies through time. Together, this suggests that existing COVID-19 studies - which have typically used cross-sectional data from the early months of the pandemic (see, for instance, Clark et al., 2020; Harper et al., 2020; Painter & Qiu, 2020) - may not be generalisable across the full pandemic. Changes in the determinants of compliance may also have implications for the targeting and phrasing of public health messaging and design of measures to maintain or improve compliance as pandemics proceed.

Therefore, in this study, we sought to test whether the association between factors relating to demographic and socio-economic characteristics, personality traits and pro-social motivations were more or less predictive of compliance with COVID-19 guidelines across five months of the pandemic in the UK. We used data from a large weekly panel study of over 21,000 adults across the period 01 April – 31 August 2020, during which time the stringency of lockdown measures in the devolved nations of the UK changed (Hale et al., 2020).

## Methods

### Participants

We used data from the COVID-19 Social Study; a large panel study of the psychological and social experiences of over 50,000 adults (aged 18+) in the UK during the COVID-19 pandemic. The study commenced on 21 March 2020 and involved online weekly data collection across the pandemic in the UK. The study is not random and therefore is not representative of the UK population, but it does contain a heterogeneous sample. The sample was recruited using three primary approaches. First, snowballing was used, including promoting the study through existing networks and mailing lists (including large databases of adults who had previously consented to be involved in health research across the UK), print and digital media coverage, and social media. Second, more targeted recruitment was undertaken focusing on (i) individuals from a low-income background, (ii) individuals with no or few educational qualifications, and (iii) individuals who were unemployed. Third, the study was promoted via partnerships with third sector organisations to vulnerable groups, including adults with pre-existing mental health conditions, older adults, carers, and people experiencing domestic violence or abuse. The study was approved by the UCL Research Ethics Committee [12467/005] and all participants gave informed consent. The study protocol and user guide (which includes full details on recruitment, retention, data cleaning, weighting and sample demographics) are available at https://github.com/UCL-BSH/CSSUserGuide.

For these analyses, we focused on participants interviewed at least once in each month between 01 April – 31 August (n = 23,252, observations = 427,161). Recruitment into the study was ongoing across this period. This sample represents 38.8% of those who were interviewed by 30 April 2020. We excluded participants with missing data on key demographic data that we use to construct survey weights (n = 627, observations = 11,480). We used complete case analysis as (a) there was only a small amount of item missingness in the study and (b) item missingness was dictated by not being interviewed during limited periods in which particular questions were included in the survey, which suggests data are missing at random.

### Lockdown measures

National lockdown was announced on 23 March 2020. Residents were required to stay at home unless purchasing essential items, exercising in nearby outdoor locations (at most once a day), or helping vulnerable individuals. Public venues were closed and businesses operated with strict social distancing guidelines or using remote working, where possible. Restrictions were gradually reduced from May 2020 with variation across the devolved nations in both the timing and extent of the reduction (Cameron-Blake et al., 2020). Domestic travel limits were removed from 13 May in England, but were relaxed later in Northern Ireland (26 June), Wales (6 July), and Scotland (7 July). Restrictions on indoor and outdoor household mixing were also gradually reduced (in June in England and Northern Ireland and July in Wales and Scotland), though local lockdowns were subsequently imposed in higher transmission areas of the UK from July. In the latter months, the UK government also began actively encouraging citizens to return to workplaces and to public venues – for instance, in August, running a subsidized meal scheme (“Eat Out to Help Out”) to support the restaurant sector. See Supplementary Figure 1.

**Figure 1:**
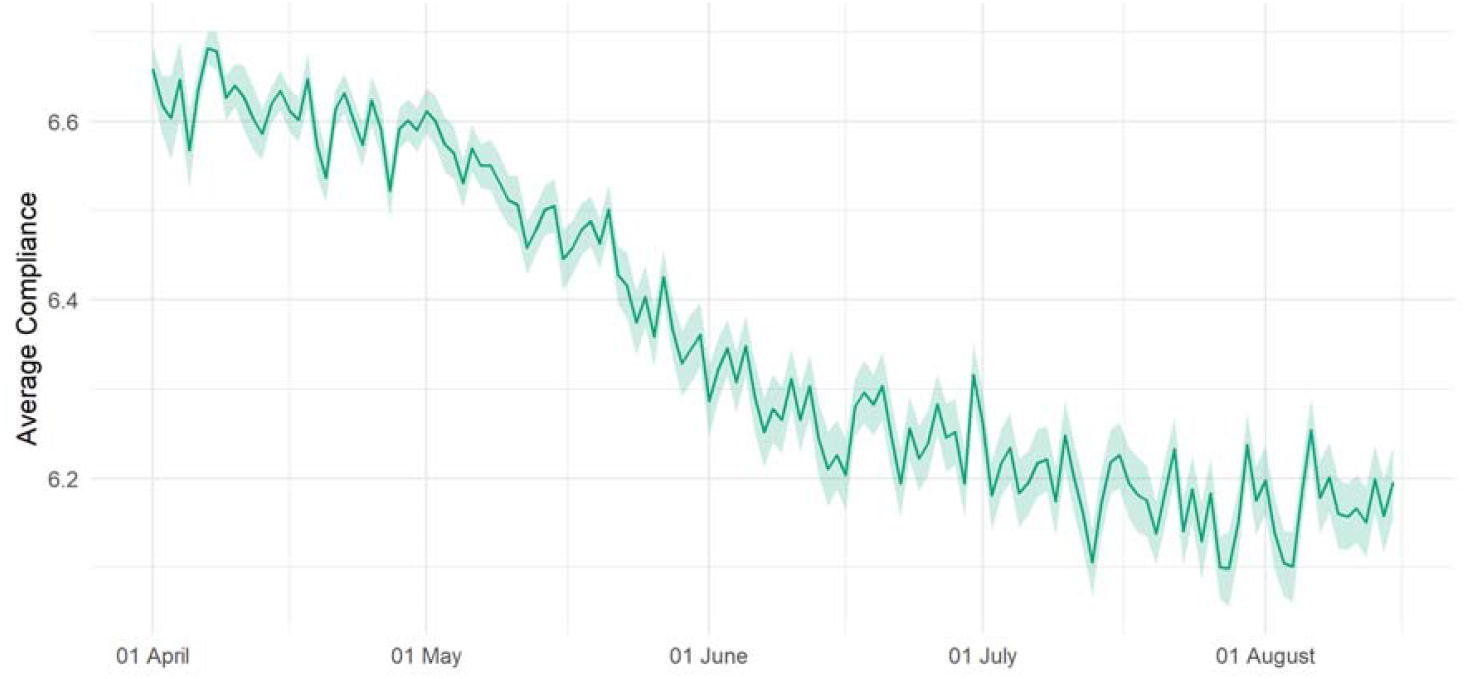
(Weighted) daily average compliance with COVID-19 guidelines, 01 April – 15 August.

### Measures

#### Compliance with COVID-19 guidelines

Compliance with guidelines was measured weekly using a single-item measure: “Are you following the recommendations from authorities to prevent spread of Covid-19?”. The item was measured on a seven-point Likert scale (1 = “Not at all”, 7 = “Very much so”), and analysed as a continuous variable.

#### Demographics and socio-economic position

We included demographic variables for country of residence (England, Scotland, Wales, Northern Ireland), sex (male or female), ethnicity (White or Non-White) and age (grouped: 18-29, 30-45, 46-59, 60+). We also included variables for socio-economic position (SEP): annual income (grouped: < £16k, £16k - £30k, £30k - £60k, £60k - £90k, £90k +), education level (higher education, further education, secondary education or below), employment status (employed, student, unemployed/inactive, retired), household overcrowding (< 1 person per room, 1+ persons per room), and living arrangement (alone, with adult but no child, with child). Each were measured at baseline interview.

#### Personality traits

Personality was measured at baseline interview using the Big Five Inventory (BFI-2; Soto & John, 2017), which measures five domains and 15 facets: *openness* (intellectual curiosity, aesthetic sensitivity, and creative imagination), *conscientiousness* (organisation, productiveness, and responsibility), *extraversion* (sociability, assertiveness, and energy level), *agreeableness* (compassion, respectfulness, and trust) and *neuroticism* (anxiety, depression, and emotional volatility). Each item was scored on a 5-point scale (1 = “strongly disagree”, 5 = “strongly agree”). We use the sum Likert score for each domain (range 3-15). As our interest is in the predictors of low compliance, in line with the results of previous studies (Brouard et al., 2020; Clark et al., 2020; Zajenkowski et al., 2020), we reverse coded items related to openness, conscientiousness, agreeableness and neuroticism so that high values indicate low levels of the trait.

*Resilience* was assessed between 14 – 21 May with the 6-item Brief Resilience Scale (Smith et al., 2008), a widely used measure of individuals’ ability to recover from stress. Items are rated on a five-point scale (1 = “strongly disagree”, 5 = “strongly agree”). We used the sum Likert score, with items coded such that higher scores indicate lower resilience (range 6 - 30).

*Locus of control* was measured between 04 – 11 June using the 6-item Locus of Control Scale developed by the University of Washington Beyond High School Project (Hirschman & Almgren, 2012), and captures generalized expectancies about whether individuals can (internal) or cannot (external) control events and outcomes in their lives. Responses were rated on a four-point scale ranging from (1 = “strongly agree”, 4 = “strongly disagree”). We used the sum score of responses, with internal worded items coded so higher values indicate more external locus of control.

*Optimism* was collected between 04 – 11 June using the 10-item Life Orientation Test-Revised (LOT-R; Scheier et al., 1994). Items are rated on a five-point scale ranging from “strongly disagree” to “strongly agree”. We used the sum Likert score, coding items such that higher scores indicate lower optimism (range 6 - 30).

*Risk-taking* was measured between 23-30 July with one item from the Dohmen Risk Taking Scale (Dohmen et al., 2011). Respondents rated the extent to which they generally see themselves as a person who is fully prepared to take risks, rated on an 11-point scale (0 = “not at all willing to take risks” to 10 = “very willing to take risks”).

#### Social and Prosocial Factors

We included several measures of pro-social motivations: empathy and social capital. *Neighbourhood social capital* before COVID-19 was measured between 09-16 July by combining four items from the social cohesion subscale of the Neighbourhood Scales (Mujahid et al., 2007) and a further item, “Before COVID-19, this was a close-knit neighbourhood”. Items were rated on a five-point scale (1 = “strongly disagree”, 5 = “strongly agree”), with items reflecting beliefs about trust, shared values, and willingness to help others. We reverse-coded items and used the sum score of items on social capital prior to COVID-19 (range 5 – 25). Higher values indicating lower social capital.

*Empathy* was assessed between 11-18 July using subscales for empathic concern and perspective-taking from the Interpersonal Reactivity Index (IRI; Davis, 1983). (Subscales for fantasy and personal distress were not administered.) *Empathic concern* (also known as emotional empathy) consists of 7 items and captures feelings of warmth, concern, and compassion for others. *Perspective-taking* (also known as cognitive empathy) assesses efforts to adopt the perspectives of others (7 items). Items were rated on a five-point scale ranging (1 = “does not describe me well”, 5 = “describes me very well”). We used the sum Likert score for each domain, coding items such that higher scores indicate lower empathy (range 0 - 28).

We also assessed four aspects of individuals’ neighbourhoods, each measured between 09-16 July. *Neighbourhood attachment* was assessed with three five-point Likert items on whether the participant feels their neighbourhood is their home (1 = “just a place to live”, 5 = “home”), their attachment to their neighbourhood (1 = “no attachment”, 5 = “strong attachment”) and feelings of belonging in their community (1 = “don’t belong at all”, 5 = “belong strongly”). We used the sum Likert score (range 3-15). We reverse-code items so that higher scores indicate lower attachment.

*Neighbourhood satisfaction* was assessed with a five-point Likert item (1 = “very dissatisfied”, 5 = “very satisfied”). We reverse code this so that higher scores indicate lower satisfaction. *Neighbourhood space* was measured using three items on satisfaction with neighbourhood walkability, usable green space, and presence of trees. Each was measure on a three-point scale (1 = “dissatisfied”, 2 = “neither satisfied nor dissatisfied”, 3 = “satisfied”), which we reverse coded and summed into a single score (range 3 – 9). *Neighbourhood crowding* was measured using three items on satisfaction with neighbourhood traffic density, noise, and levels of crowding. Each was measured on a three-point scale (categories as above), which we reverse coded and summed into a single score (range 3 – 9).

### Statistical Analysis

Our basic empirical strategy was to estimate multilevel models of the form:

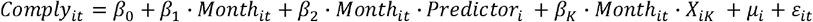

Where *i* is an index of individuals and *t* is an index of time. *μ*_*i*_ is a person-specific random error (i.e. a random intercept) and *ε*_*it*_ is an observation-specific random error, both normally distributed. *Month*_*it*_ is a categorical indicator of the month the interview was recorded in, *Predictor*_*i*_ is a specific predictor under study, and *X*_*iK*_ is a set of control variables (defined below). Our interest is therefore in the coefficient *β*_2_, which shows how the association between the predictor and compliance differs across time.

For each predictor variable, we estimated two models: an unadjusted model with no control variables (except month), and an adjusted model that included (i) factors we identified as core confounders (including demographic and socio-economic variables as defined above, pre-existing psychiatric diagnosis, and Big-5 personality traits) and (ii) factors likely to account for differences in compliance behaviours (including self or family member “shielding” at any point due to pre-existing health conditions, whether the participant was remaining indoors for other pre-existing health reasons e.g. physical disability, and the number of long-term physical health conditions (categorical: 0, 1, 2+). We used complete case data and survey weights when estimating models. Derivation of the survey weights is described in further detail in the Supplementary Information. For comparability, we report standardized coefficients.

An issue with restricting the analysis to individuals who were interviewed each month is that results may be biased by non-random attrition from the survey. In particular, individuals who report high compliance with COVID-19 guidelines are less likely to drop out (see Supplementary Information Figure S2), which may bias towards finding smaller associations. Consequently, as a sensitivity analysis, we repeat models using data from balanced panels with fewer months of data.

## Results

### Descriptive Statistics

Descriptive statistics for the sample are displayed in **Error! Reference source not found**.. There was a significant decline in compliance with COVID-19 guidelines across the study period (Figure 1). When comparing descriptive statistics and compliance levels according to last month of interview in the study, participants who were included in the main analysis (i.e. interviewed in August 2020) had higher and less steep drops in compliance, were older, less likely to be employed, and differed on several personality traits, including higher optimism and resilience and lower neuroticism and openness to experience (Table S1 and Supplementary Figure S2). Repeated cross-sectional data from YouGov (2020) on the performance of specific preventative behaviours showed declines in avoiding crowds and social gatherings between April and August, while mask-wearing and hand-washing increased (Supplementary Figure S3).

**Table 1:**
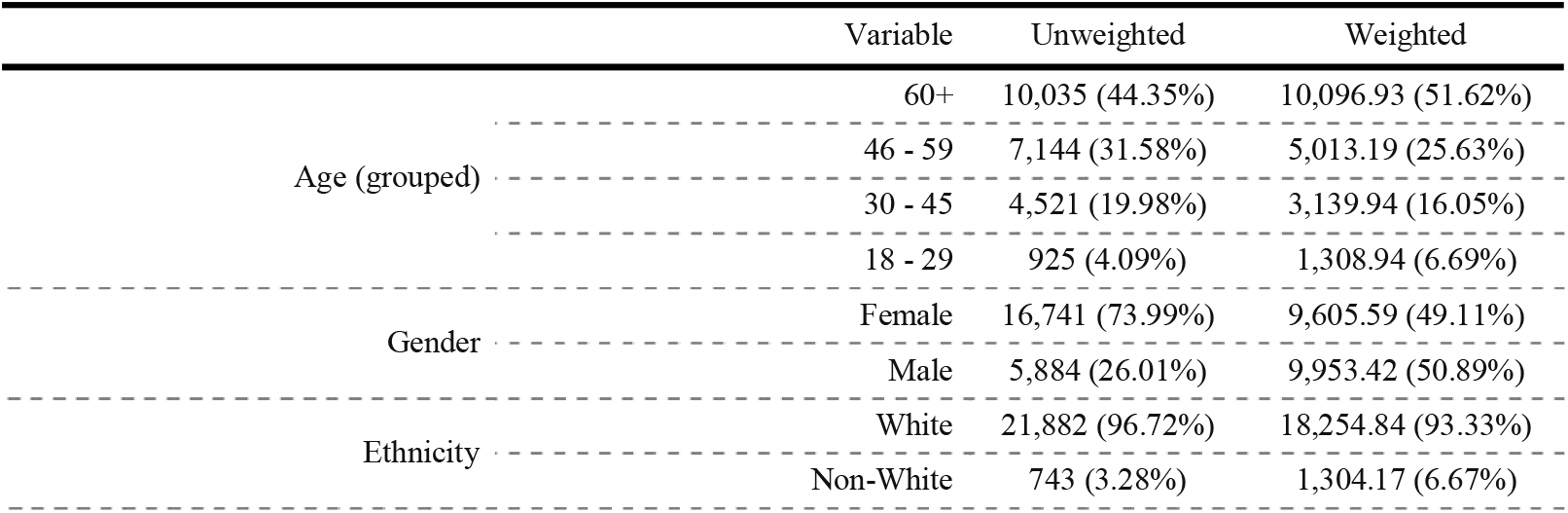
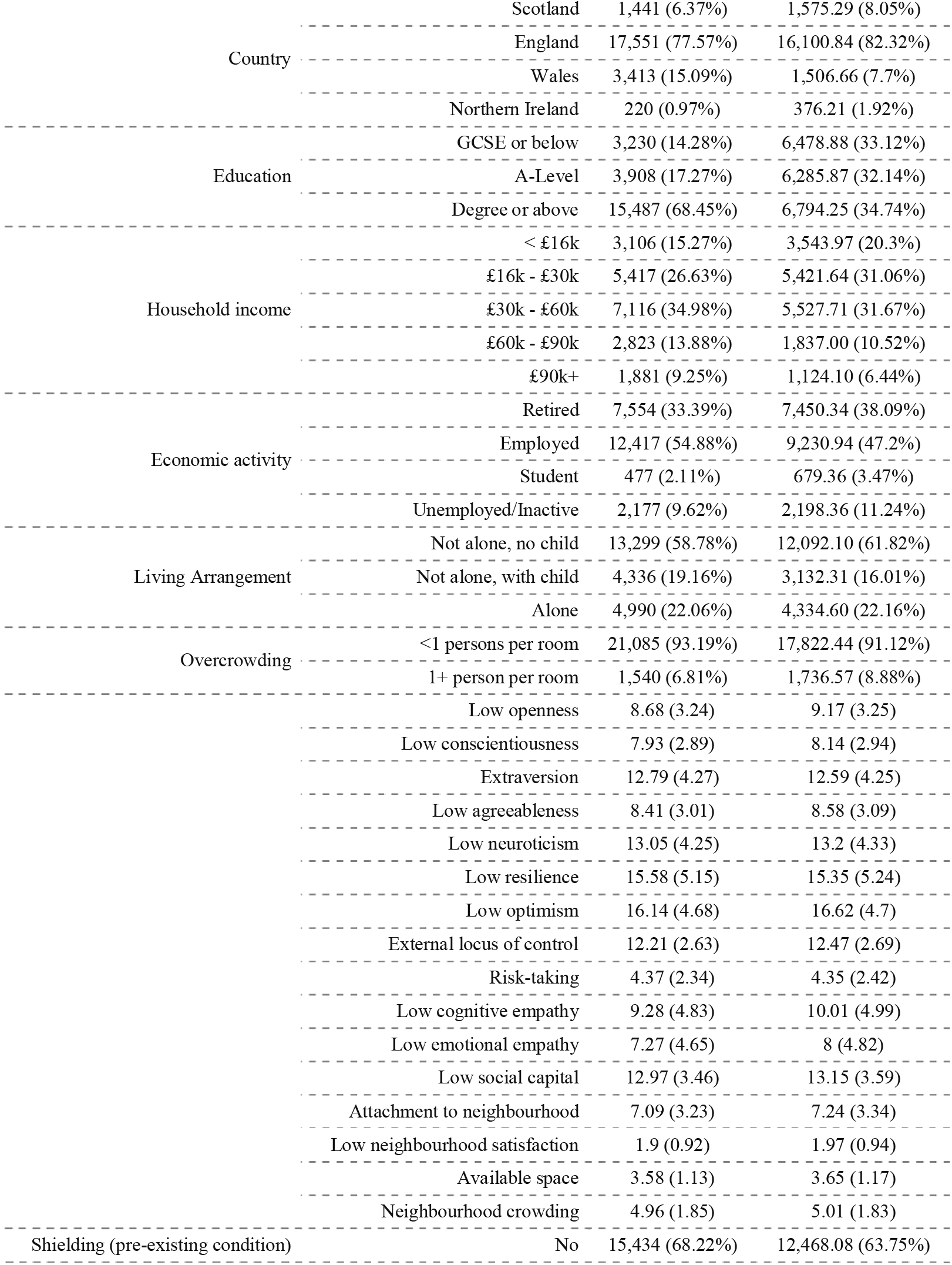
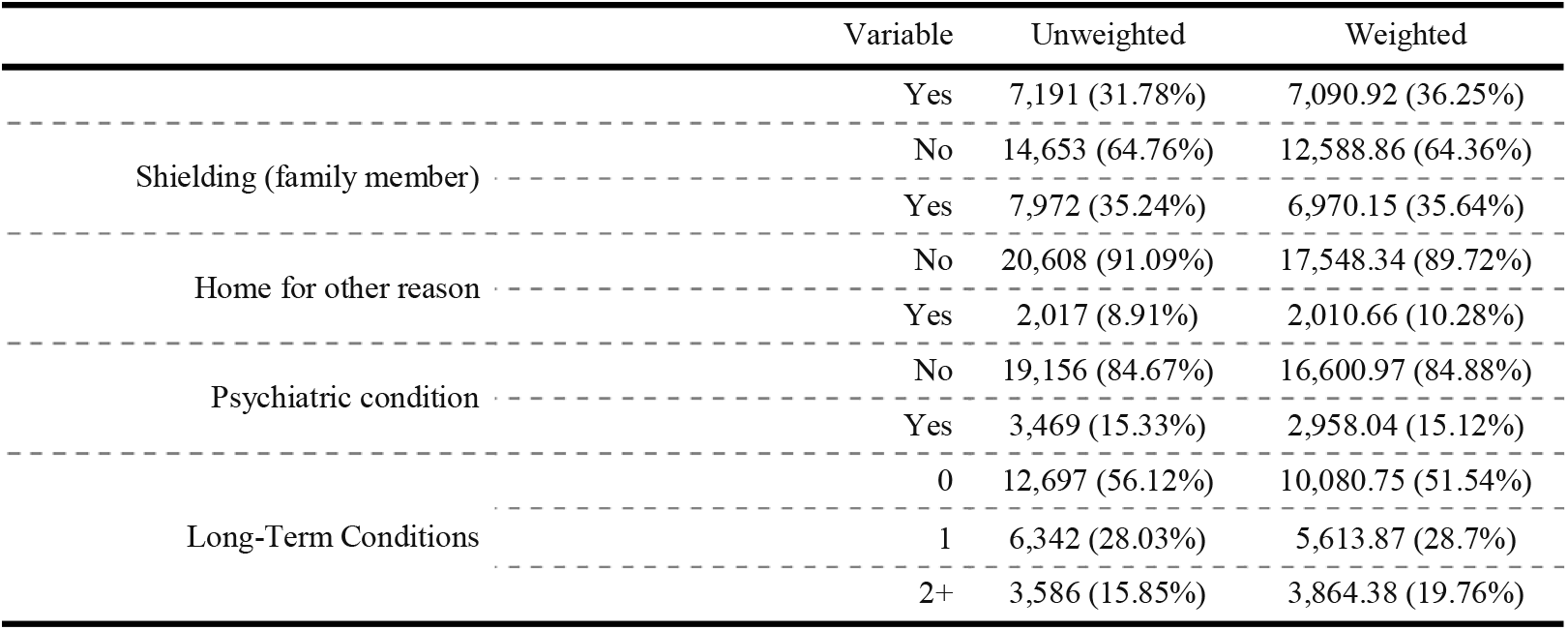
Descriptive Statistics

### Demographics and Socio-Economic Position

Age-group, employment status, county of residence, income group, gender, living arrangement and household overcrowding were each related to compliance in multivariate models (Figure 2; bivariate results, which are substantively similar, are shown in Supplementary Figure S4.) Younger adults had lower levels of compliance than older adults in April and these age-related differences grew considerably as the pandemic continued. Females reported higher compliance than males, though there was little difference by month. There was little difference by ethnic group, but there was evidence that compliance was higher in Scotland than in England, to a somewhat greater extent in August than April/May. There was also an association between higher education and higher household income and lower compliance that grew across time. Students and individuals in employment had lower compliance levels than retired individuals. Comparing students and retirees, the difference was larger in later months. Adults living with a child or in overcrowded accommodation had lower compliance rates, with this increasing over the summer months. Effect sizes were generally small (< 0.25 SD), except for age in latter months: by August, 18-29 year olds had 0.72 SD (95% CI = 0.79, 0.66) lower compliance than participants aged 60+.

**Figure 2:**
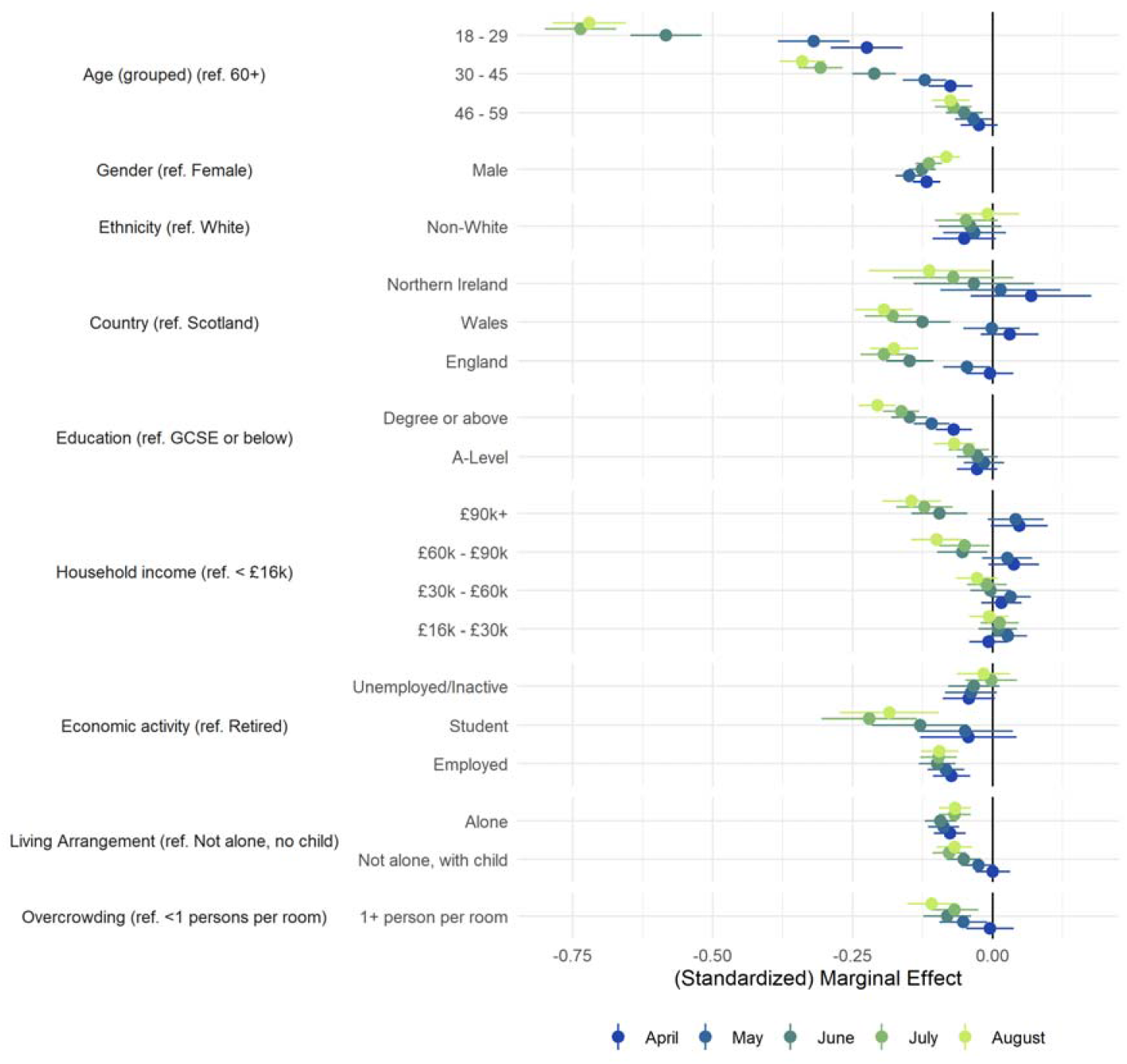
Association between socio-economic and demographic characteristics and compliance with COVID-19 guidelines by month, derived from mixed effects models. Models include adjustment for sex, age group, ethnicity, education, income group, employment status, country of residence, living arrangement, household overcrowding, whether the participant is shielding, diagnosed psychiatric condition, long-term physical health conditions, and Big-5 personality traits

### Personality traits

Each of the Big-5 traits, resilience, pessimism, locus of control, and risk tacking were related to compliance (Figure 3; note the different scale used for the x-axis). The strongest association was with risk taking, for which the association was almost twice as large in July/August than in April. Conscientiousness, openness to experience, and extraversion also become more highly related to compliance in later months, which pessimism became less strongly associated. Associations between compliance and agreeableness, resilience, locus of control and neuroticism were little changed. The bivariate results are substantively similar, except for extraversion, which was related to higher compliance (Supplementary Figure S5). Effect sizes were small (less than approximately 0.2 SD), in each case.

**Figure 3:**
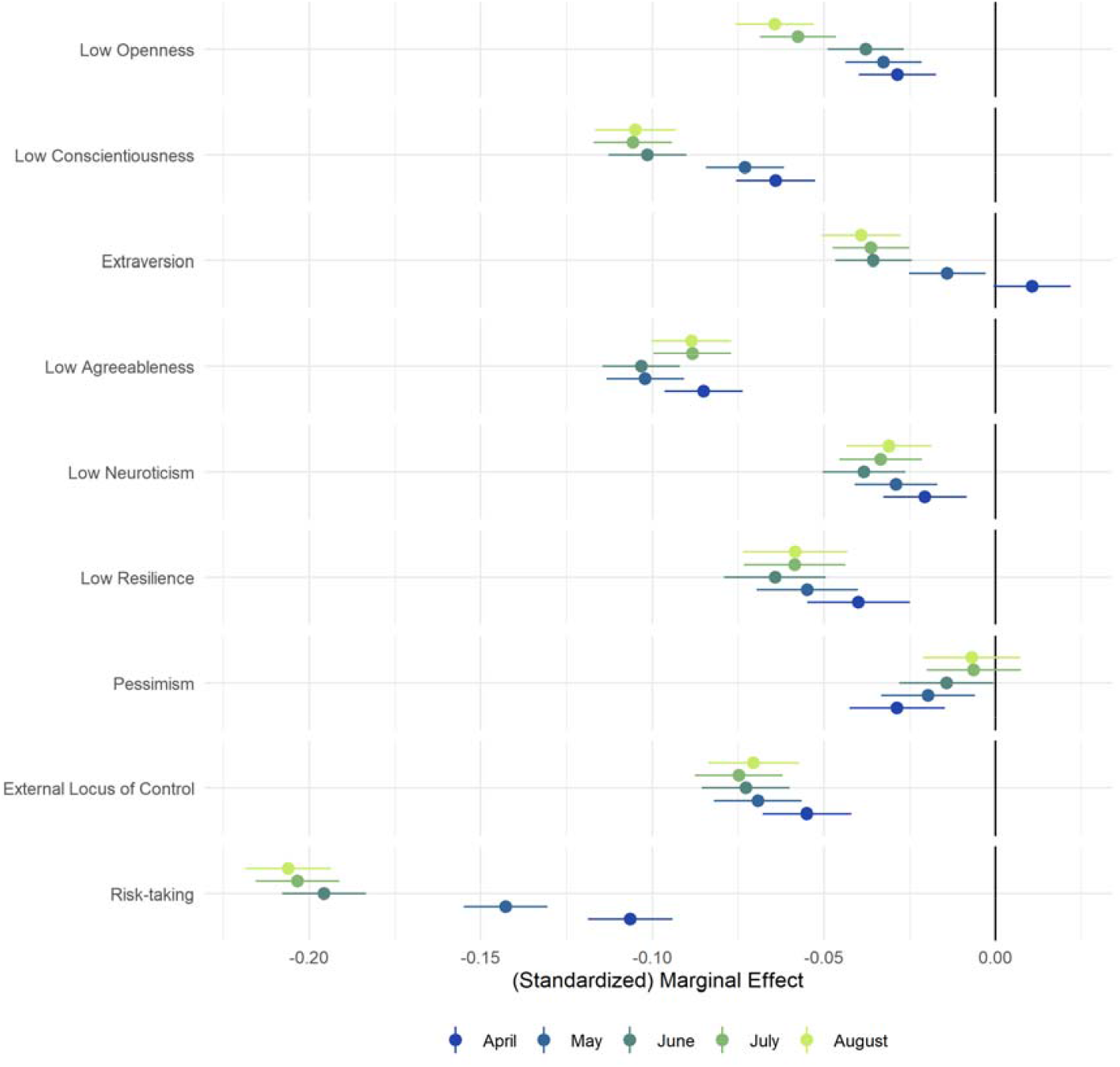
Association between personality traits and compliance with COVID-19 guidelines by month, derived from mixed effects models. Left panel: bivariate association. Models include adjustment for sex, age group, ethnicity, education, income group, employment status, country of residence, living arrangement, household overcrowding, whether the participant is shielding, diagnosed psychiatric condition, long-term physical health conditions, and Big-5 personality traits.

### Social and Prosocial Factors

Lower empathy, neighbourhood attachment and social capital were each related to lower compliance (Figure 4). People with higher levels of cognitive or emotional empathy were more likely to continue complying in later months, while there was little change in associations for the other factors. Dissatisfaction with neighbourhood, available space and neighbourhood crowding were each related to lower compliance. The association between neighbourhood characteristics and compliance was broadly stable across time. Bivariate results are substantively similar, with the exception that there was no clear increase in the size of the association between low empathy and compliance in later months (Supplementary Figure S6). Effect sizes were small in each instance.

**Figure 4:**
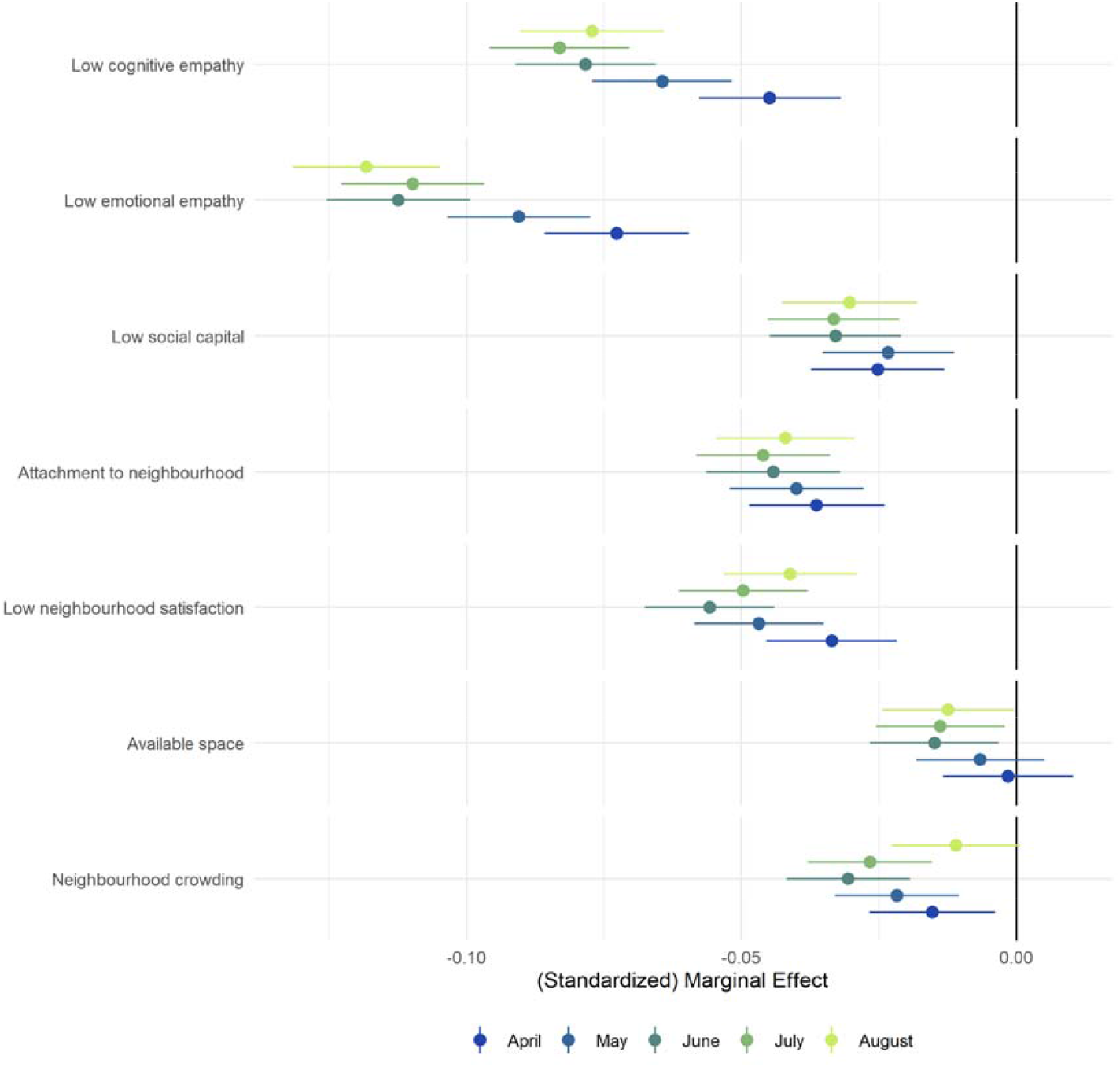
Association between prosocial motivations, neighbourhood factors and compliance with COVID-19 guidelines by month, derived from mixed effects models. Models include adjustment for sex, age group, ethnicity, education, income group, employment status, country of residence, living arrangement, household overcrowding, whether the participant is shielding, diagnosed psychiatric condition, long-term physical health conditions, and Big-5 personality traits.

### Sensitivity Analysis

Results were qualitatively similar using balanced panels who dropped out before August (Supplementary Figures S7-S8).

## Discussion

This study explored changes in the between-person predictors of compliance with COVID-19 guidelines in a longitudinal sample of UK adults across the first five months of social distancing and lockdown measures in the UK. Compliance was associated with major demographic and socio-economic characteristics, personality traits, prosocial motivations and neighbourhood characteristics. In general, effect sizes were small, with the exception of a strong association in later months with age. Specifically, decreases in average compliance levels over the period examined were more pronounced among young people (see Supplementary Figure S9), individuals low in conscientiousness and empathic concern, and individuals with risk-seeking attitudes. However, associations with compliance did not differ over the study period for all characteristics, with some (including agreeableness, neighbourhood attachment and neighbourhood characteristics) not changing in strength as predictors of compliance over time.

Some of the predictors of compliance identified in this study have been identified in previous studies, including low empathy, age, gender, and social capital (Bish & Michie, 2010; Jørgensen et al., 2020; Zajenkowski et al., 2020). The finding that demographic characteristics (besides age) are not strongly related to compliance is also consistent with previous studies in the literature, which generally show that major individual demographic and socioeconomic characteristics and personality traits do not explain much of the between-person variation in compliance levels (see, for instance, Brouard et al., 2020; Clark et al., 2020; Zajenkowski et al., 2020). However, previous literature has been cross-sectional and marred with inconsistencies in findings, and the results from this study help to explain why. Instead of being constant predictors of compliance over time, the differential strength of so many predictors of compliance over the five months of the study suggests that the determinants of compliance are context-specific. One explanation for these contextual differences is “situational strength”, whereby behaviour is less determined by personal characteristics in contexts where options are constrained and/or normative behaviour is clearly prescribed (Cooper & Withey, 2009; Götz et al., 2020; Zajenkowski et al., 2020). In support of this theory, decreases in average compliance coincided with lockdown and social distancing restrictions becoming less stringent. Indeed, we found that compliance decreased fastest in England and Wales (where restrictions were eased fastest) compared to Scotland (where restrictions were kept more stringent for longer) (Cameron-Blake et al., 2020). A further potential explanation for the differential strength of predictors is that as restrictions continued for longer, boredom increased and self-control depleted, meant that individuals’ abilities to over-ride their own individual traits and adhere to restrictive rules decreased and the true predictive power of these predictors became more evident (Bieleke et al., 2020; Martarelli & Wolff, 2020). Therefore, this study highlights the importance of considering individual traits as evolving predictors of compliance and, in particular, considering the role of context as a moderator of the relationship between traits and compliance.

The results have a number of policy implications. First, the findings suggest that compliance with pandemic control measures decreases as the stringency of measures is reduced. This highlights the importance of reinforcing messaging on compliance as measures are eased to avoid perceptions that remaining measures are somehow unnecessary. Second, the results suggest some individuals may not be responsive to specific types of communications. For example, individuals with high risk-taking propensities may be less responsive to messages about personal risk but more responsive to prosocial messages about the impact of their risk-taking on others. Therefore, in seeking to maximize compliance amongst different groups, a plurality of communication approaches is required. It also highlights why punitive measures for non-compliance may not be especially effective (Kooistra et al., 2020; Murphy et al., 2020). For example, in September 2020, the UK government increased fines for those caught violating household mixing rules. Whilst such measures have the potential to reduce non-compliance among those for whom personal risk of infection or protecting others is not sufficient motivation, it risks reducing pro-social motivations (Gneezy & Rustichini, 2000) and highlighting the extent of norm violations, which, for some, could reduce compliance. Third, the finding that individuals living in overcrowded accommodation and neighbourhoods with little space had lower and faster decreasing compliance suggests that poor quality housing and crowded lived environments could exacerbate challenges for governments in tackling public health emergencies, over and above the greater risk of interpersonal transmission due to higher proximity (Rader et al., 2020). As such, tackling social inequalities has consequences not only for general public health but also for behavioural management during pandemics.

That said, we also found that individuals with higher incomes had higher initial compliance but faster decreases over time. It is possible that these individuals were able to maintain a strict compliance initially due to not facing any financial barriers such as an inability to pay bills that may have driven to rules being broken in a search for work. However, as the pandemic continued, greater wealth and a sense of privilege or a lack of financial fear over fines may have driven a more relaxed approach to compliance. Given research showing that the non-compliance of people in positions of power has a negative impact on societal trust and others’ compliance (Fancourt et al., 2020), this highlights the importance of the consistent application of pandemic rules amongst all groups.

This study had a number of limitations. Although we included a wide array of demographic and personality variables in our models, results may still have been explained by unobserved confounding. However, given that the strength of some of the factors studied here differed across the pandemic (e.g. risk taking), it is questionable how confounding factors could have generated these temporal patterns. Second, participants self-reported their compliance with measures. Results may, therefore, have been biased due to social desirability concerns and due to differences in knowledge. People’s understanding of compliance may also have differed over time, particular as rules changed. We also lack detail on the specific rules – such as mask-wearing, social distancing or hand-washing – that individuals were breaking when reporting lower compliance. Some of the factors studied here may be important for some behaviours rather than others – for instance, extraversion is more plausibly related to violating social distancing rules than non-mask wearing. So future studies are encouraged that explore specific compliance behaviours in more detail. A final issue is the use of a non-random sample. While the sample was heterogeneous and we included population weights in models, the data were from a study set-up explicitly to research COVID-19. It is likely that individuals who participated (and continued to participate) in the study had a higher interest in helping tackle the pandemic than the general population at large. This interest may manifest as a higher propensity to comply with guidelines. Further, selection biases are likely to have arisen due to different modes of recruitment into the survey. Therefore, the relationships identified here may be biased estimates of the actual predictive effects of some of these factors.

Nevertheless, this study demonstrated that the importance of many factors in predicting compliance has differed across the pandemic and is therefore context specific. Further, the results are in line with theories of “situational strength” where individual characteristics are more important where behaviour is less constrained. This highlights the need to account for context when studying compliance and to study predictors of compliance as evolving factors. For policy makers and public health professionals, the results suggest that if we want to maintain good compliance or increase compliance, two key things are needed. First, multiple messages are required to target different groups given capabilities and motivations will vary substantially between different demographic groups. Second, these messages will need to evolve across pandemics as the context and behavioural opportunities for individuals change.

## Data Availability

Data from the COVID-19 Social Study will be made public at the end of the pandemic. The code to replicate the analysis is available at https://osf.io/e2c6g.

## Supplementary Material

**Figure S1:**
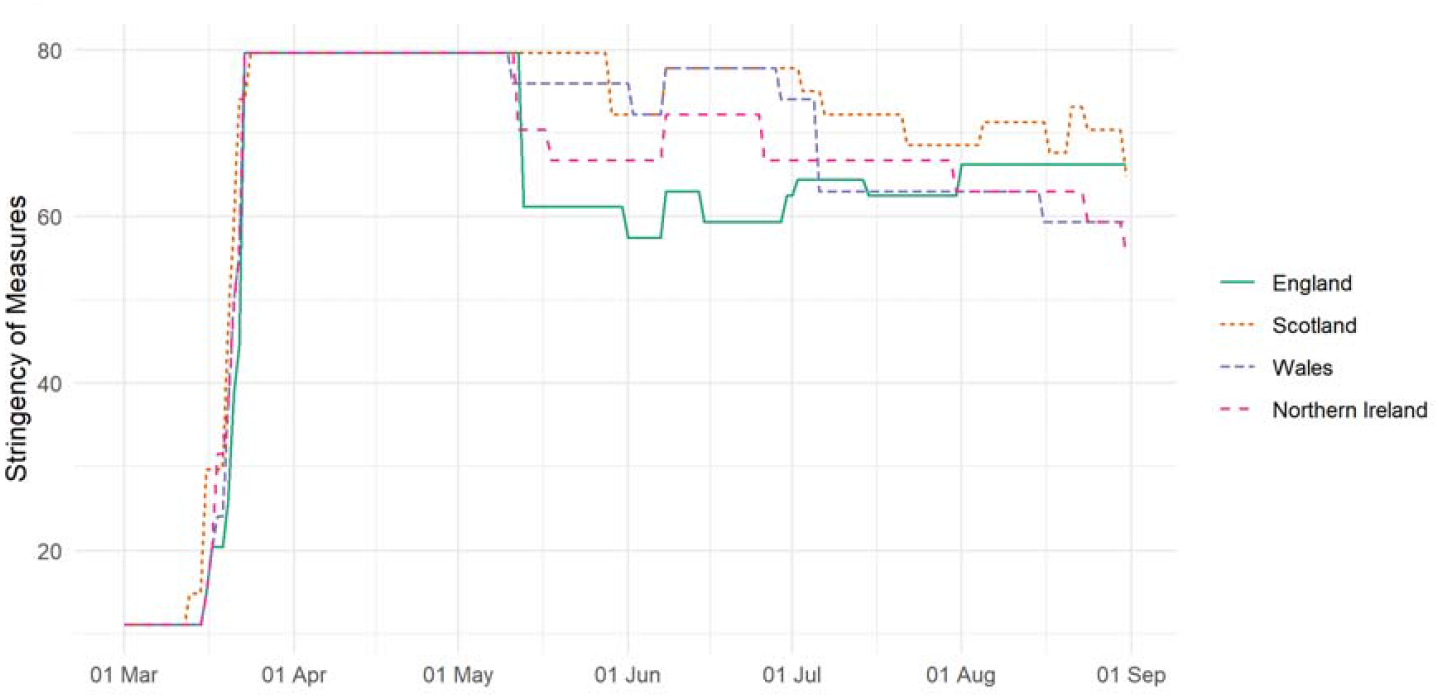
Stringency of UK government COVID-19 related measure, 02 January – 31 July. Source: Oxford COVID-19 Government Response Tracker (Hale et al., 2020).

**Figure S2:**
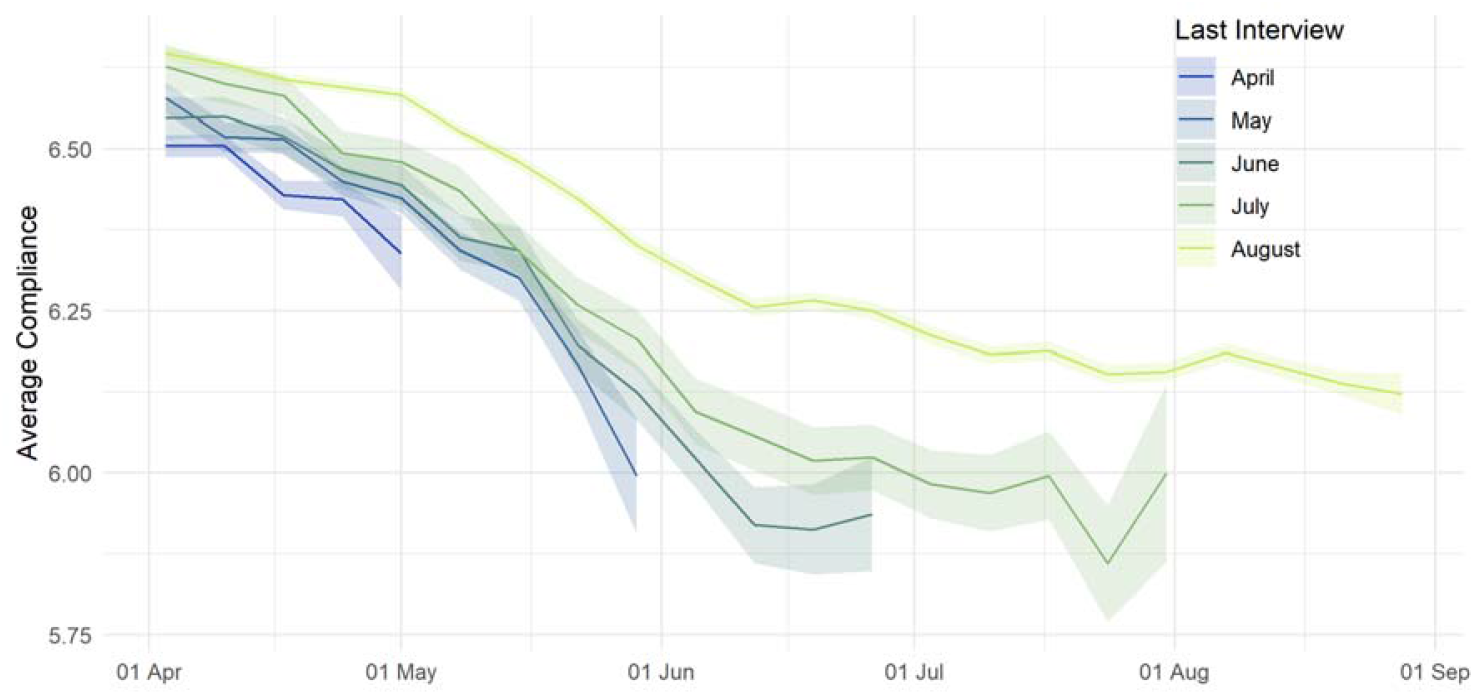
(Weighted) weekly average compliance by month of last interview. Participants with last interview in August were included in the main analysis.

**Figure S3:**
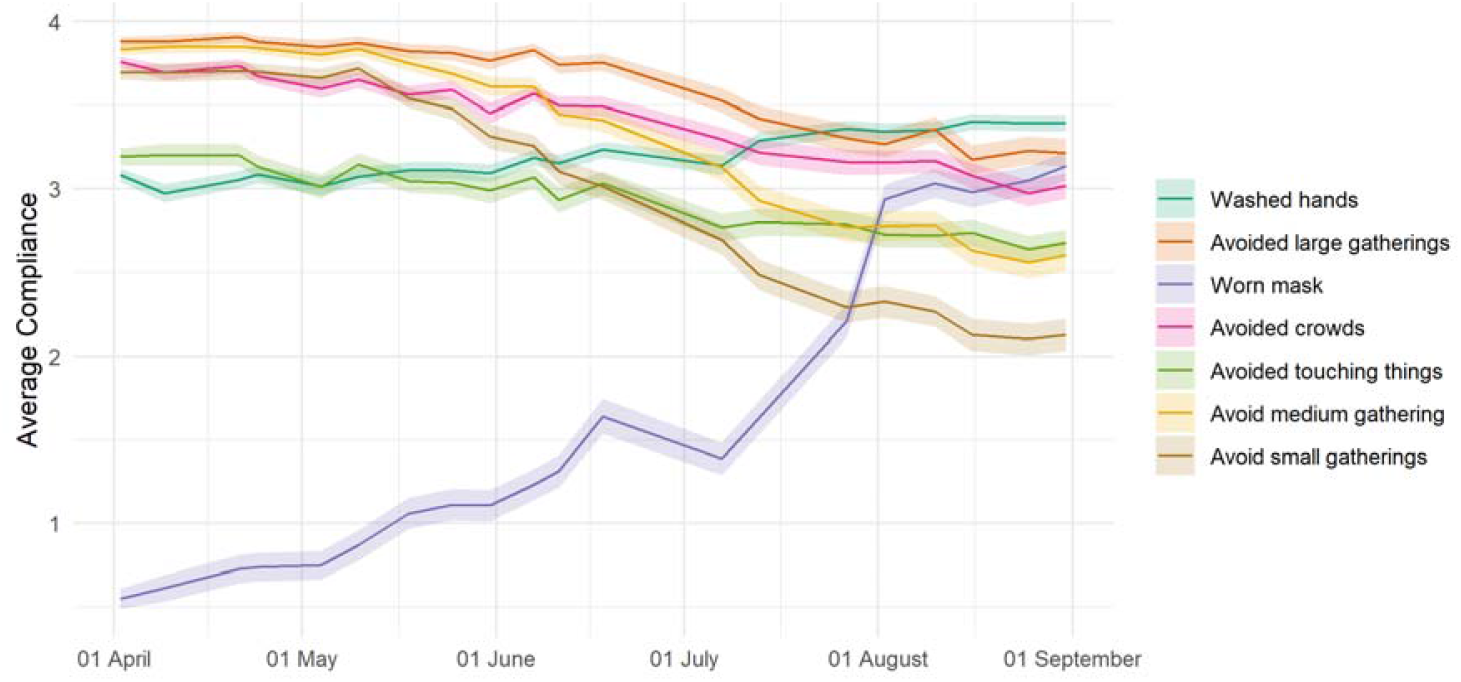
(Weighted) weekly average compliance with recommended preventative behaviours in the UK, 01 April – 31 August. Source: YouGov COVID-19 Public Monitor (YouGov, 2020).

**Figure S4:**
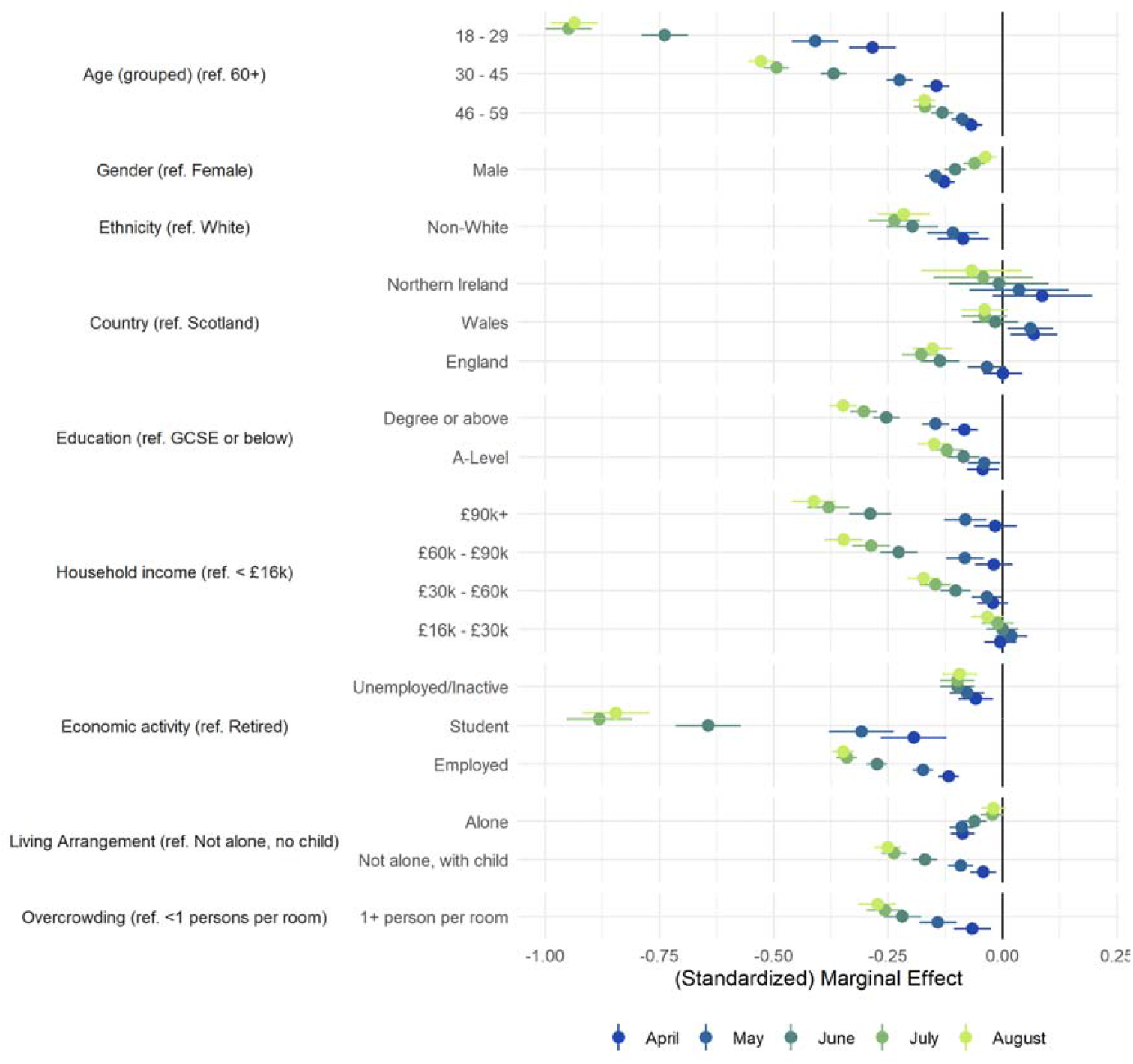
Association between demographic and socio-economic characteristics and compliance with COVID-19 guidelines by month, derived from mixed effects models. Bivariate associations.

**Figure S5:**
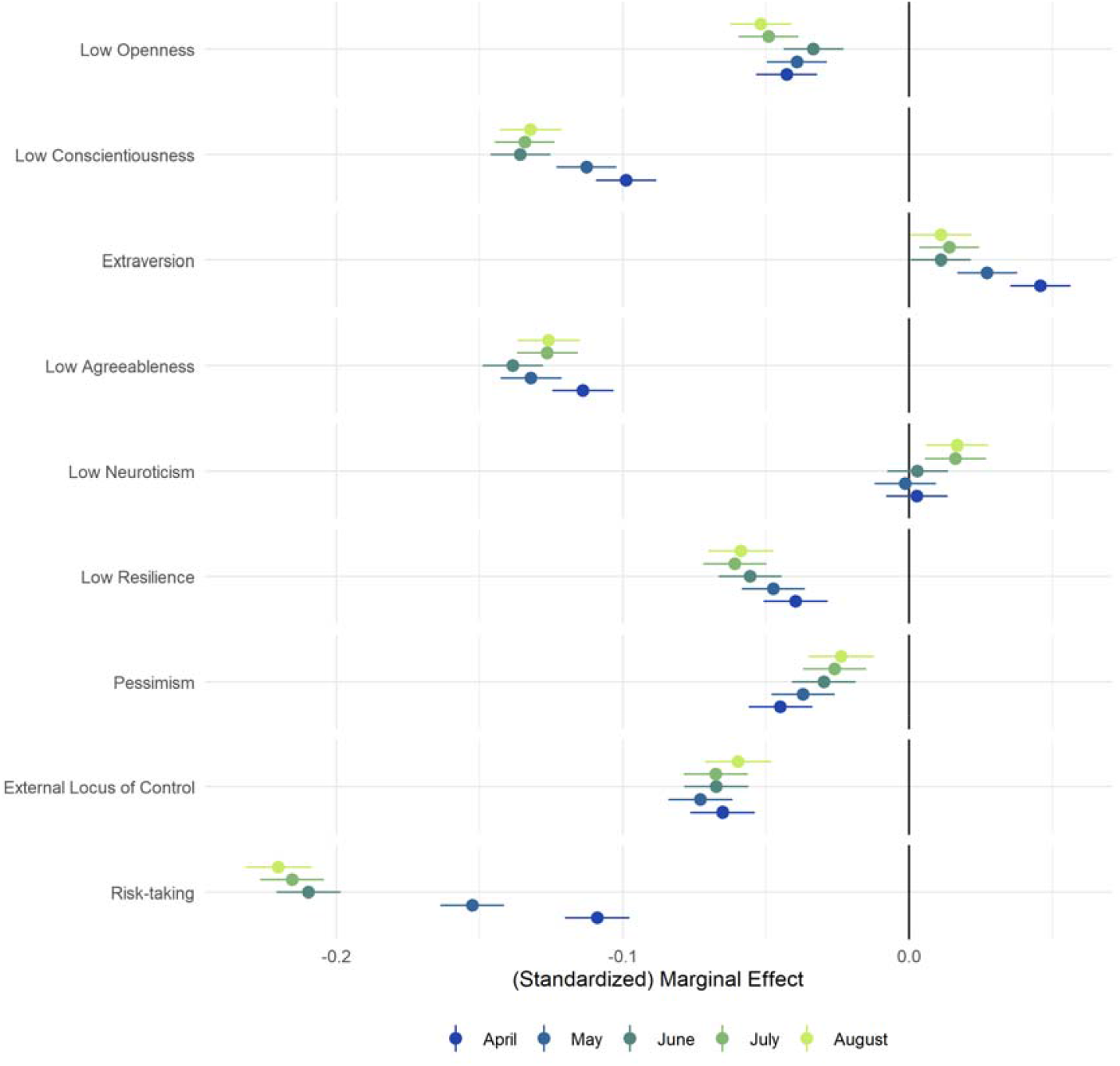
Association between personality traits and compliance with COVID-19 guidelines by month, derived from mixed effects models. Bivariate associations.

**Figure S6:**
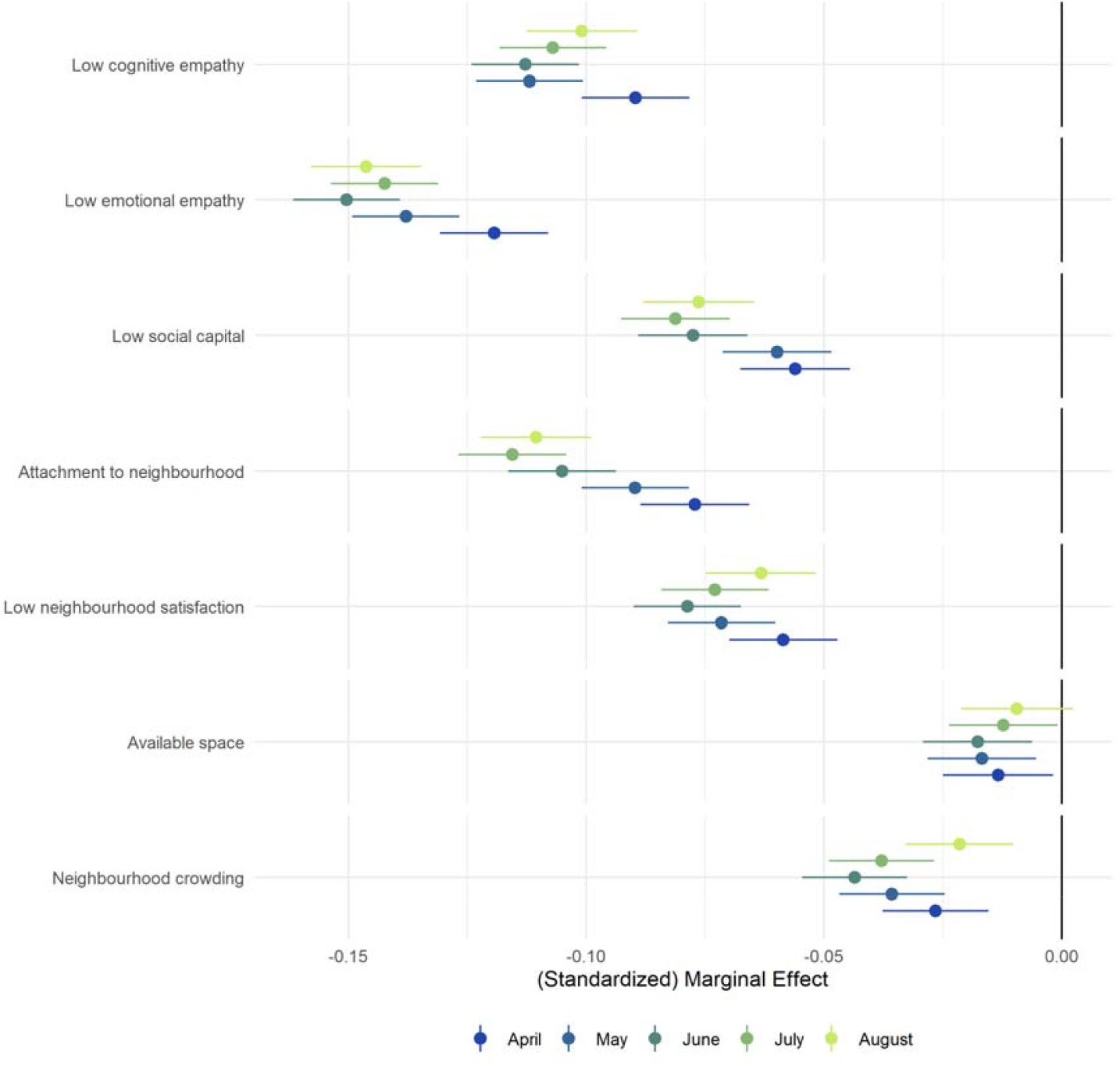
Association between social and pro-social factors and compliance with COVID-19 guidelines by month, derived from mixed effects models. Bivariate associations.

**Figure S7:**
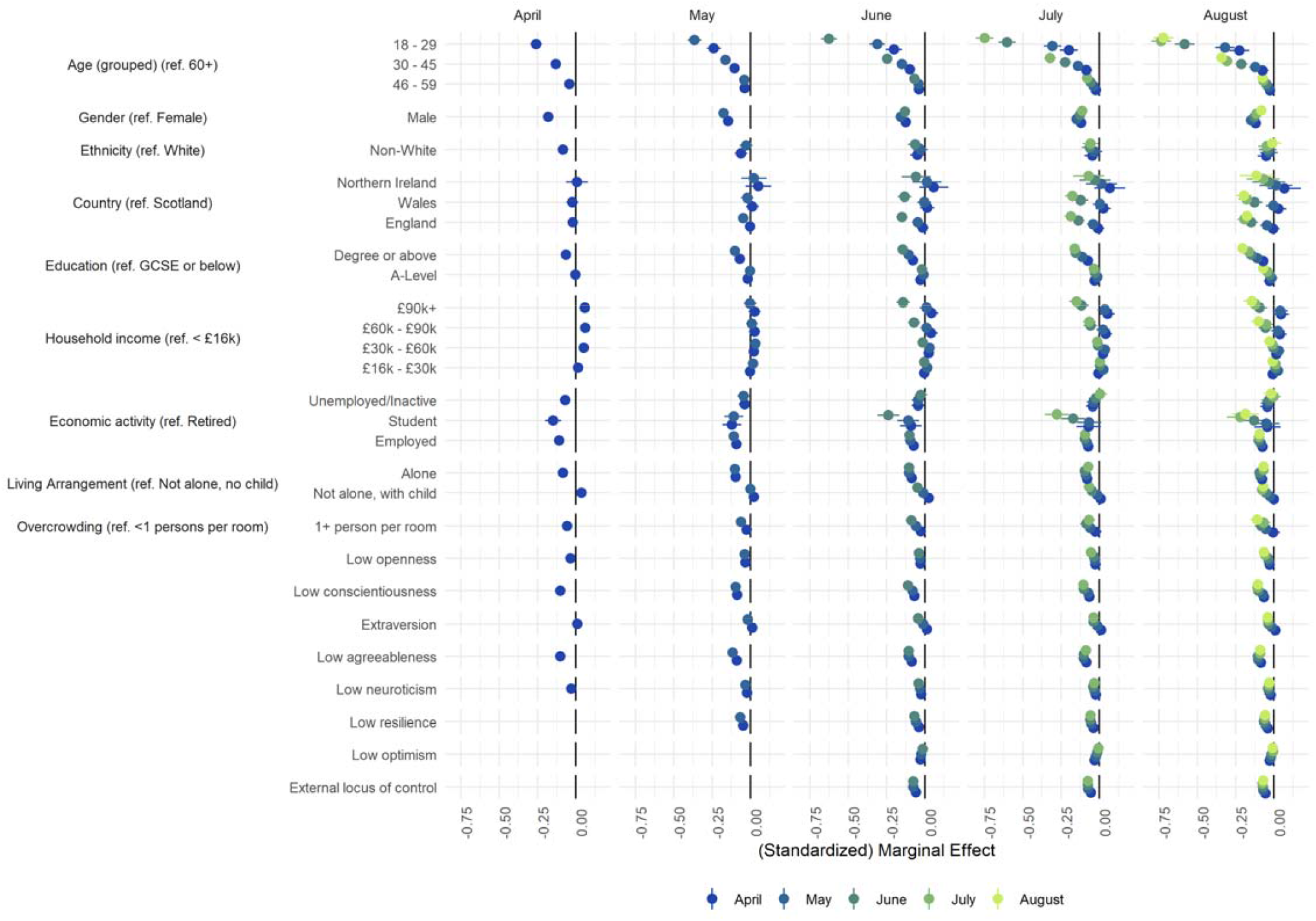
Association between demographic and socio-economic characteristics, personality traits and social and pro-social factors and compliance with COVID-19 guidelines by month, derived from mixed effects models. Each panel represents a difference balance panel of individuals interviewed up to a particular month. Models include adjustment for sex, age group, education, income group, employment status, country of residence, living arrangement, household overcrowding, whether the participant is shielding, diagnosed psychiatric condition, and Big-5 personality traits.

**Figure S8:**
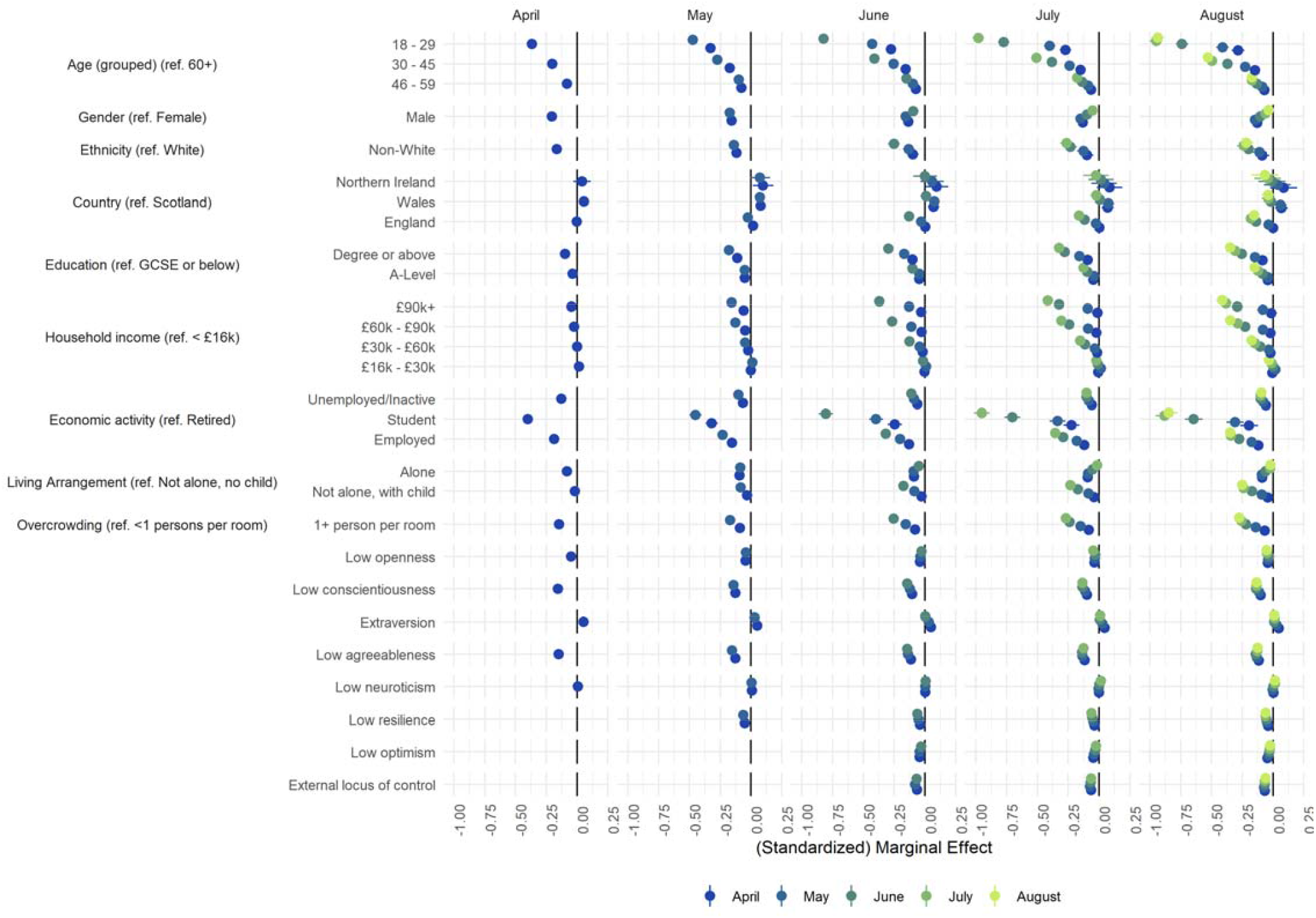
Association between demographic and socio-economic characteristics, personality traits and social and pro-social factors and compliance with COVID-19 guidelines by month, derived from mixed effects models. Each panel represents a difference balance panel of individuals interviewed up to a particular month. Bivariate associations.

**Figure S9:**
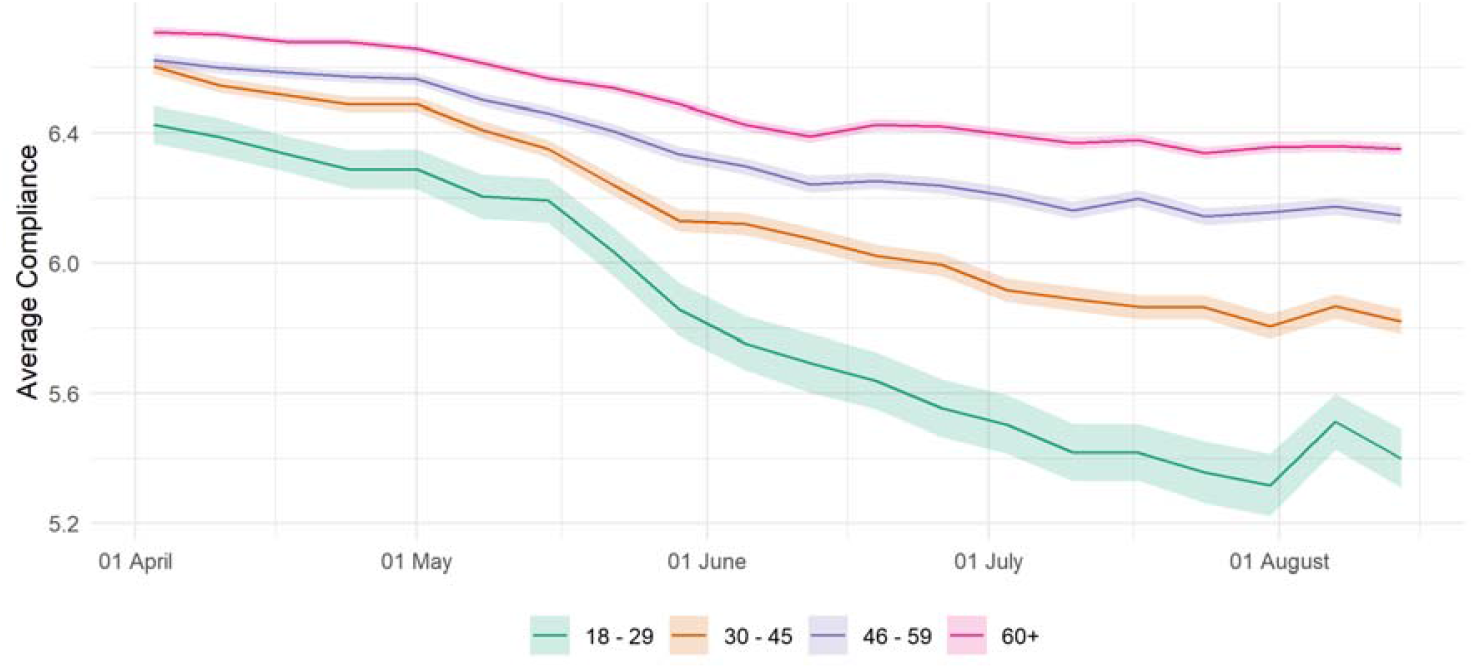
(Weighted) weekly average compliance by age group.

**Table S1:**
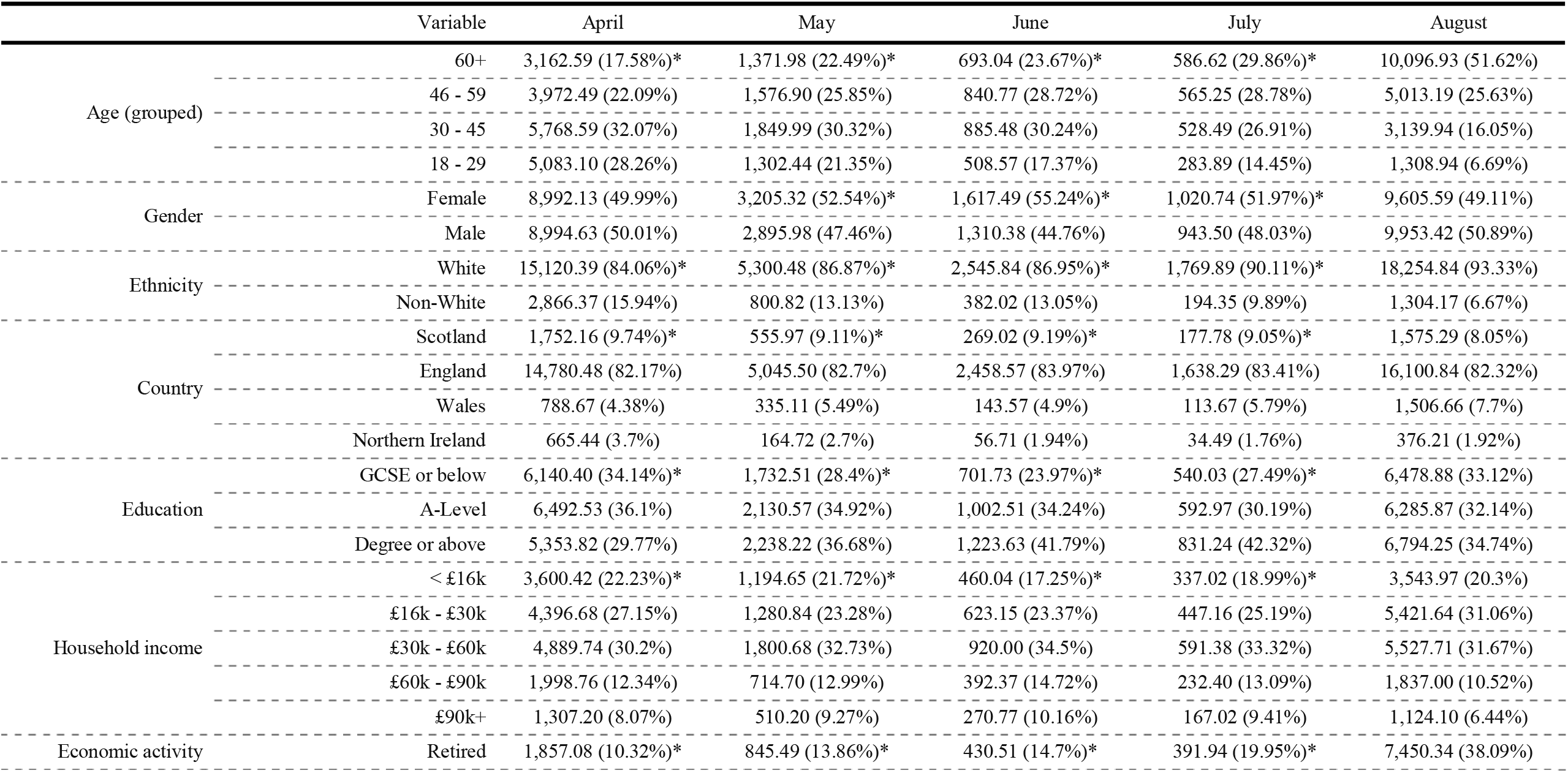
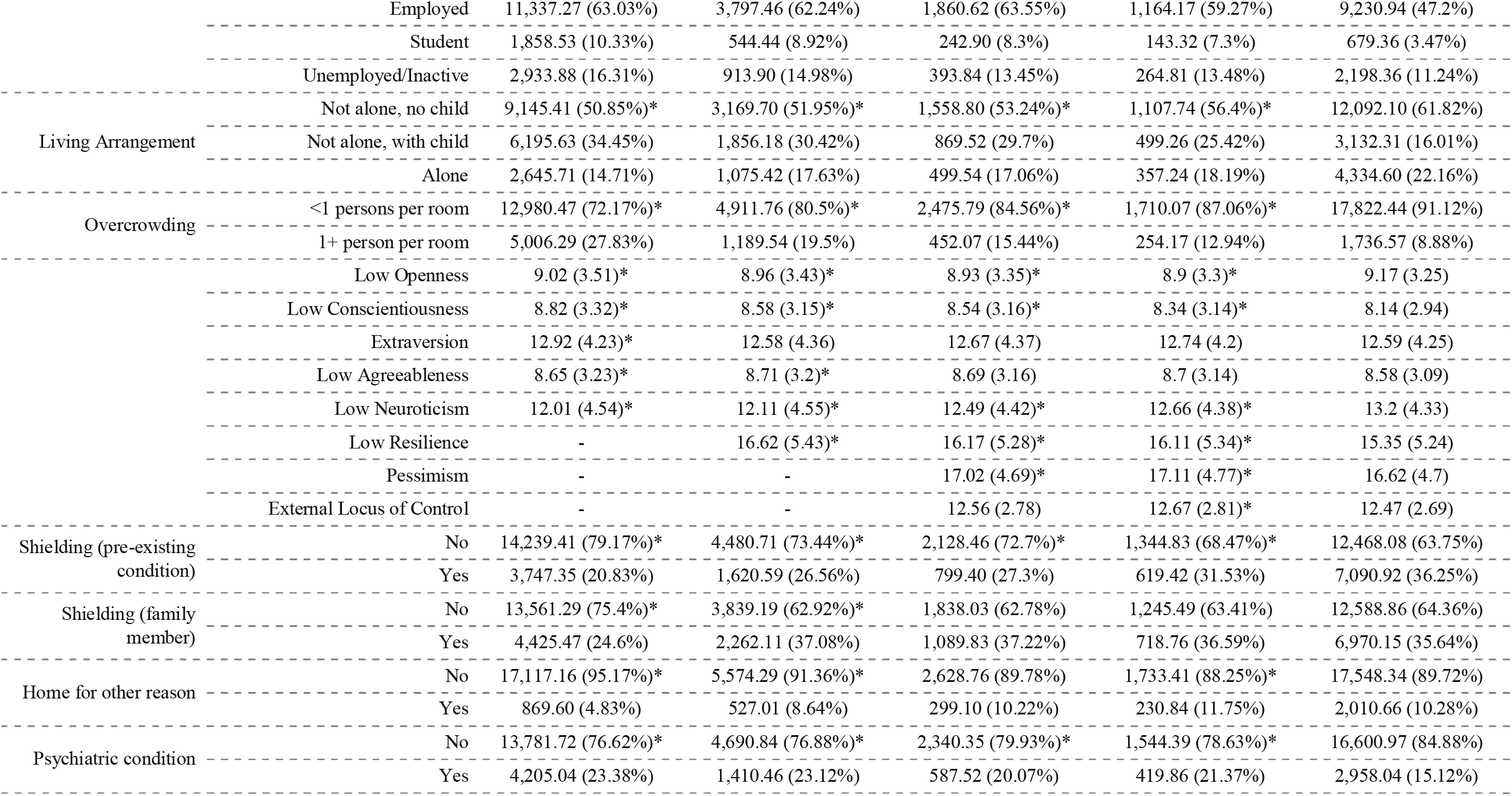
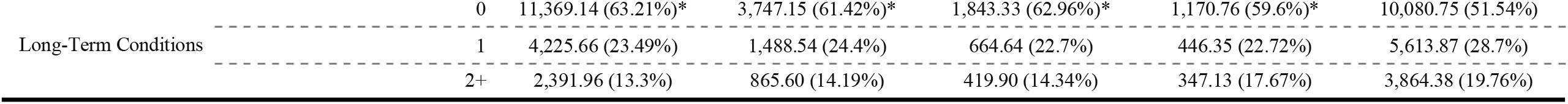
(Weighted) descriptive statistics by last month of interview. Participants interviewed in July are included in the main analysis. Participants last interviewed in earlier months are excluded.

## References

Adams-Prassl, A., Boneva, T., Golin, M., & Rauh, C. (2020). Inequality in the impact of the coronavirus shock: Evidence from real time surveys. Journal of Public Economics, 189, 104245. https://doi.org/10.1016/j.jpubeco.2020.104245

Bieleke, M., Martarelli, C., & Wolff, W. (2020). Boredom makes it difficult, but it helps to have a plan: Investigating adherence to social distancing guidelines during the COVID-19 pandemic [Preprint]. PsyArXiv. https://doi.org/10.31234/osf.io/enzbv

Bish, A., & Michie, S. (2010). Demographic and attitudinal determinants of protective behaviours during a pandemic: A review. British Journal of Health Psychology, 15(4), 797–824. https://doi.org/10.1348/135910710X485826

Brouard, S., Vasilopoulos, P., & Becher, M. (2020). Sociodemographic and Psychological Correlates of Compliance with the COVID-19 Public Health Measures in France. Canadian Journal of Political Science, 1–6. https://doi.org/10.1017/S0008423920000335

Cameron-Blake, E., Tatlow, H., Wood, A., Hale, T., Kira, B., Petherick, A., & Phillips, T. (2020). Variation in the response to COVID-19 across the four nations of the United Kingdom (Working Paper BSG-WP-2020/035; BSG Working Papers). Blavatnik School of Government, University of Oxford. https://www.bsg.ox.ac.uk/research/publications/variation-response-covid-19-across-four-nations-united-kingdom

Chu, D. K., Akl, E. A., Duda, S., Solo, K., Yaacoub, S., Schünemann, H. J., Chu, D. K., Akl, E. A., El-harakeh, A., Bognanni, A., Lotfi, T., Loeb, M., Hajizadeh, A., Bak, A., Izcovich, A., Cuello-Garcia, C. A., Chen, C., Harris, D. J., Borowiack, E., … Schünemann, H. J. (2020). Physical distancing, face masks, and eye protection to prevent person-to-person transmission of SARS-CoV-2 and COVID-19: A systematic review and meta-analysis. The Lancet, 395(10242), 1973–1987. https://doi.org/10.1016/S0140-6736(20)31142-9

Clark, C., Davila, A., Regis, M., & Kraus, S. (2020). Predictors of COVID-19 voluntary compliance behaviors: An international investigation. Global Transitions, 2, 76–82. https://doi.org/10.1016/j.glt.2020.06.003

Cooper, W. H., & Withey, M. J. (2009). The Strong Situation Hypothesis. Personality and Social Psychology Review, 13(1), 62–72. https://doi.org/10.1177/1088868308329378

Cowling, B. J., Ng, D. M. W., Ip, D. K. M., Liao, Q., Lam, W. W. T., Wu, J. T., Lau, J. T. F., Griffiths, S. M., & Fielding, R. (2010). Community Psychological and Behavioral Responses through the First Wave of the 2009 Influenza A(H1N1) Pandemic in Hong Kong. The Journal of Infectious Diseases, 202(6), 867–876. https://doi.org/10.1086/655811

Davis, M. H. (1983). Measuring individual differences in empathy: Evidence for a multidimensional approach. Journal of Personality and Social Psychology, 44(1), 113–126. https://doi.org/10.1037/0022-3514.44.1.113

Dohmen, T., Falk, A., Huffman, D., Sunde, U., Schupp, J., & Wagner, G. G. (2011). INDIVIDUAL RISK ATTITUDES: MEASUREMENT, DETERMINANTS, AND BEHAVIORAL CONSEQUENCES. Journal of the European Economic Association, 9(3), 522–550. https://doi.org/10.1111/j.1542-4774.2011.01015.x

Fancourt, D., Steptoe, A., & Wright, L. (2020). The Cummings effect: Politics, trust, and behaviours during the COVID-19 pandemic. The Lancet, 396(10249), 464–465. https://doi.org/10.1016/S0140-6736(20)31690-1

Gneezy, U., & Rustichini, A. (2000). A Fine Is a Price. The Journal of Legal Studies, 29(1), 1–17. https://doi.org/10.1086/468061

Götz, F. M., Gvirtz, A., Galinsky, A. D., & Jachimowicz, J. M. (2020). How personality and policy predict pandemic behavior: Understanding sheltering-in-place in 55 countries at the onset of COVID-19. American Psychologist. https://doi.org/10.1037/amp0000740

Hale, T., Angrist, N., Cameron-Blake, E., Hallas, L., Kira, B., Majumdar, S., Petherick, A., Phillips, T., Tatlow, H., & Webster, S. (2020). Oxford COVID-19 Government Response Tracker. Blavatnik School of Government. https://www.bsg.ox.ac.uk/research/research-projects/coronavirus-government-response-tracker

Harper, C. A., Satchell, L. P., Fido, D., & Latzman, R. D. (2020). Functional Fear Predicts Public Health Compliance in the COVID-19 Pandemic. International Journal of Mental Health and Addiction. https://doi.org/10.1007/s11469-020-00281-5

Hirschman, C., & Almgren, G. (2012). University of Washington - Beyond High School (UW-BHS): Version 5 [Data set]. ICPSR - Interuniversity Consortium for Political and Social Research. https://doi.org/10.3886/ICPSR33321.V5

Jørgensen, F. J., Bor, A., & Petersen, M. B. (2020). Compliance Without Fear: Individual-Level Predictors of Protective Behavior During the First Wave of the COVID-19 Pandemic [Preprint]. PsyArXiv. https://doi.org/10.31234/osf.io/uzwgf

Kooistra, E. B., Folmer, C. R., Kuiper, M. E., Olthuis, E., Brownlee, M., Fine, A., & Rooij, B. van. (2020). Mitigating COVID-19 in a Nationally Representative UK Sample: Personal Abilities and Obligation to Obey the Law Shape Compliance with Mitigation Measures. PsyArXiv. https://doi.org/10.31234/osf.io/zuc23

Martarelli, C. S., & Wolff, W. (2020). Too bored to bother? Boredom as a potential threat to the efficacy of pandemic containment measures. Humanities and Social Sciences Communications, 7(1), 1–5. https://doi.org/10.1057/s41599-020-0512-6

Michie, S., van Stralen, M. M., & West, R. (2011). The behaviour change wheel: A new method for characterising and designing behaviour change interventions. Implementation Science, 6(1), 42. https://doi.org/10.1186/1748-5908-6-42

Mongey, S., Pilossoph, L., & Weinberg, A. (2020). Which Workers Bear the Burden of Social Distancing Policies? (No. w27085; p. w27085). National Bureau of Economic Research. https://doi.org/10.3386/w27085

Mujahid, M. S., Diez Roux, A. V., Morenoff, J. D., & Raghunathan, T. (2007). Assessing the Measurement Properties of Neighborhood Scales: From Psychometrics to Ecometrics. American Journal of Epidemiology, 165(8), 858–867. https://doi.org/10.1093/aje/kwm040

Murphy, K., Williamson, H., Sargeant, E., & McCarthy, M. (2020). Why people comply with COVID-19 social distancing restrictions: Self-interest or duty? Australian & New Zealand Journal of Criminology, 0004865820954484. https://doi.org/10.1177/0004865820954484

Painter, M., & Qiu, T. (2020). Political Beliefs affect Compliance with COVID-19 Social Distancing Orders. SSRN Electronic Journal. https://doi.org/10.2139/ssrn.3569098

Park, C. L., Russell, B. S., Fendrich, M., Finkelstein-Fox, L., Hutchison, M., & Becker, J. (2020). Americans’ COVID-19 Stress, Coping, and Adherence to CDC Guidelines. Journal of General Internal Medicine, 35(8), 2296–2303. https://doi.org/10.1007/s11606-020-05898-9

Rader, B., Scarpino, S. V., Nande, A., Hill, A. L., Adlam, B., Reiner, R. C., Pigott, D. M., Gutierrez, B., Zarebski, A. E., Shrestha, M., Brownstein, J. S., Castro, M. C., Dye, C., Tian, H., Pybus, O. G., & Kraemer, M. U. G. (2020). Crowding and the shape of COVID-19 epidemics. Nature Medicine, 1–6. https://doi.org/10.1038/s41591-020-1104-0

Scheier, M. F., Carver, C. S., & Bridges, M. W. (1994). Distinguishing optimism from neuroticism (and trait anxiety, self-mastery, and self-esteem): A reevaluation of the Life Orientation Test. Journal of Personality and Social Psychology, 67(6), 1063–1078. https://doi.org/10.1037/0022-3514.67.6.1063

Smith, B. W., Dalen, J., Wiggins, K., Tooley, E., Christopher, P., & Bernard, J. (2008). The brief resilience scale: Assessing the ability to bounce back. International Journal of Behavioral Medicine, 15(3), 194–200. https://doi.org/10.1080/10705500802222972

Soto, C. J., & John, O. P. (2017). The next Big Five Inventory (BFI-2): Developing and assessing a hierarchical model with 15 facets to enhance bandwidth, fidelity, and predictive power. Journal of Personality and Social Psychology, 113(1), 117–143. https://doi.org/10.1037/pspp0000096

van der Weerd, W., Timmermans, D. R., Beaujean, D. J., Oudhoff, J., & van Steenbergen, J. E. (2011). Monitoring the level of government trust, risk perception and intention of the general public to adopt protective measures during the influenza A (H1N1) pandemic in the Netherlands. BMC Public Health, 11(1), 575. https://doi.org/10.1186/1471-2458-11-575

Wang, C., Pan, R., Wan, X., Tan, Y., Xu, L., Ho, C. S., & Ho, R. C. (2020). Immediate Psychological Responses and Associated Factors during the Initial Stage of the 2019 Coronavirus Disease (COVID-19) Epidemic among the General Population in China. International Journal of Environmental Research and Public Health,17(5), 1729. https://doi.org/10.3390/ijerph17051729

Webster, R. K., Brooks, S. K., Smith, L. E., Woodland, L., Wessely, S., & Rubin, G. J. (2020). How to improve adherence with quarantine: Rapid review of the evidence. Public Health, 182, 163–169. https://doi.org/10.1016/j.puhe.2020.03.007

YouGov. (2020, October 5). Personal measures taken to avoid COVID-19. https://yougov.co.uk/topics/international/articles-reports/2020/03/17/personal-measures-taken-avoid-covid-19

Zajenkowski, M., Jonason, P. K., Leniarska, M., & Kozakiewicz, Z. (2020). Who complies with the restrictions to reduce the spread of COVID-19?: Personality and perceptions of the COVID- 19 situation. Personality and Individual Differences,166,110199. https://doi.org/10.1016/j.paid.2020.110199

